# Child health, nutrition and gut microbiota development during the first two years of life; study protocol of a prospective cohort study from the Khyber Pakhtunkhwa, Pakistan

**DOI:** 10.1101/2024.10.23.24315915

**Authors:** Muhammad Shahzad, Muhammad Ismail, Benjamin Misselwitz, Ahsan Saidal, Simon C Andrews, Khalid Iqbal, Hatice Akarsu Egger, Ziad Al Nabhani

## Abstract

Recent evidence suggests that gut microbiota development during infancy impact several metabolic, immune and endocrine pathways in humans. An imbalance in the gut microbiota diversity or function, also known dysbiosis, not only affect early child growth and development, but also linked with the development of chronic, non-communicable diseases in later life. The CHAMP (**C**hild **H**ealth **A**nd **M**icrobiome develpment study – **P**akistan) study aims to longitudinally assess gut microbiota development and associated factors (maternal, child and demographic) during early childhood, in populations residing in malnutrition endemic communities in Pakistan. A prospective cohort of mother-infant pairs (n=70) will be recruited from District Swat, Pakistan and followed for two years. Complete information about demographic characteristics, anti-natal and post-natal care, dietary intake and feeding practices and child health will be collected at baseline and 3, 6, 12, 18 and 24 months. Anthropometric measurements (height, weight, mid upper arm circumference and head circumference), dry blood spot and fecal samples will also be collected. Ethical approval of the study has been obtained from Khyber Medical University Pakistan. The study is also registered on clincaltrial.gov (Ref no: NCT05793294). The study finding will help researchers understand gut microbiota development, associated factors and its impact on longitudinal growth in infants during the first two years of life.

## Introduction

Infancy, the period lasting from birth to around two years of age is a crucial stage in human development characterized primarily by rapid physical, mental and emotional growth (1). During this period, many environmental factors such as gestational age, gender, ethnicity, dietary intake and nutritional status and socioeconomic conditions significantly affect child growth, development and overall health (2,3). However, until know, these factors either alone or synergistically, have failed to explain the overall spectrum and variations in children. Recently, research studies have emphasized that apart from intersection of environmental factors, the gut microbiome also plays an important role in mediating growth and development especially during infancy.

The human fetus is believed to develop in a microbe free environment (4). However, immediately after birth, the infant gut especially the distal ileum and colon is rapidly colonized by different microorganisms (bacteria, fungi, archaea and viruses) by vertical transmission from maternal body sites such as vagina, skin, oral cavity and gut (5). In newborn infants, gut microbiota exhibits low diversity consisting mainly of facultative anaerobic bacteria such as *Proteobacteria* (e.g., *Escherichia* and *Enterobacter*) and *Firmicutes* (*Staphylococcus, Enterococcus*and *Streptococcus*) (6). The early colonization has a profound influence on gut environment and subsequent colonization by obligate anaerobes such as *Bifidobacterium, Bacteroides* and *Clostridium*. The relative abundance of *Bifidobacteria* gradually increases 3 – 4 days after birth and become a predominant bacterial genus when the baby is around 1 month old (7,8). In healthy growing children, the gut microbiota gradually increases in complexity with highly diverse microbial composition during and after the first two to three years of life (9). During this period, the patterned ecological assembly of microbes mainly occurs under the influence of delivery mode, type of feeding, complimentary feeding practices, antibiotic use and geography (9–12). Gut microbiota development during early life impacts several metabolic, immune and endocrine pathways and is thus intimately linked to child growth and development (13). Therefore, any disturbances either in the host-microbe coevolution, microbial composition or functional potential of gut microbiome commonly referred to as microbial dysbiosis can impair immune functions, growth and development in children.

In recent years, several studies have reported associations between gut microbiome dysbiosis and impaired growth and development in children from different countries across the word. As an examples, a causal relationship between gut microbiome immaturity, undernutrition and impaired growth has been reported in 6- and 18-months old children (14). Similarly, a Swedish cohort encompassing 471 children has also reported significantly lower microbial diversity and reduced abundance of *Faecalibacterium* and *Ruminococcus* in the gut microbiome of children with slower weight gain compared with normal (15). Research studies from developing countries, where malnutrition is common, have also reported impaired or immature gut microbiome in malnourished (stunting, wasting and underweight) children compared to healthy controls. In Bangladesh, children with severe acute malnutrition (SAM) exhibit a considerably less diverse gut microbiome, low relative abundance of *Bacteroidetes* and high relative abundance of *Proteobacteria* especially the pathogen/pathobiont genera such as *Klebsiella, Escherichia, Shigella*, and *Streptococcus* (16). In another study, a group of SAM infants aged 6-to 24-months, the absolute abundance of *B. infantis* was considerably low compared to age-matched healthy infants (17). These findings were also confirmed by research studies conducted in other developing countries from Asia and Africa (18–20). Altogether, these findings suggest that an immature/poorly developed gut microbiome in infants and young children is associated with malnutrition and its consequences especially, impaired growth and development. As a result, a nutrition intervention strategy that does not take into account gut microbiome will fail to ameliorate long term consequences of malnutrition including child health and cognitive development (21). In fact, clinical studies in animals and humans have already demonstrated a positive impact of microbiome based, nutritional interventions on gut microbiome and ponderal growth (22,23). These findings may have important implications for low- and middle-income countries like Pakistan where malnutrition in children is a crucial public health challenge.

Pakistan is the 5^th^ most populous country in the world (approx. 250 million population) (24)with enormous economic, geopolitical, social and poverty-related challenges (25). According to the latest national nutrition survey, the prevalence of stunting (40.2%), wasting (17.7%) and micronutrient deficiencies in children under five years of age (26) remained at unacceptable high levels. For the last five years, the country is placed in the “Serious” category of the Global Hunger Index (27). The consequences of malnutrition are also catastrophic economically with $7.6 billion/annum (∼3% of GDP) lost to malnutrition (28). Over the last two decades, there has been virtually no improvement in the overall nutritional status of the Pakistani population despite efforts from the government and developmental organizations. Without coordinated efforts, it is very unlikely that Pakistan can achieve the UN Sustainable Development Goals to end hunger and all forms of malnutrition by 2030 (29). Successful tackling of the issue requires cost-effective, culturally relevant and sustainable nutritional interventions. Keeping in view the central role of the gut microbiome mediating malnutrition in children, it is crucial to map the compositional and functional dynamics of the gut microbiome and associated maternal, child and environment related factors during early childhood, in populations residing in malnutrition endemic communities in Pakistan. Therefore, the current study attempts to characterize how the gut microbiome develops in a cohort of children at high risk of malnutrition in Khyber Pakhtunkhwa province of Pakistan.

## Objectives

The primary objective of the CHAMP study is to characterize gut microbiota development and longitudinal growth in infants during the first two years of life. This will be done by assessing the microbial diversity, relative abundance of dominant microbiota and interindividual variations in microbial diversity and infant growth patterns. The secondary objective of the study is to investigate the impact of (i) sociodemographic characteristics (ii) mode of delivery (iii) maternal factors such diet, anti-natal and postnatal care (iv) child dietary intake and feeding practices and (v) nutritional status of the child on gut microbiota composition at different time points from birth until the child is two years old.

### Operational definitions

- **Nutritional status:** Nutritional status of the child will be measured in terms of weight for age (WAZ), height for age (HAZ), and weight for height (WHZ) according to WHO reference range (30).
- **Underweight:** Defined as weight for age of the child below two standard deviations (−2SD) from the median weight for age of the reference population.
- **Stunting:** Defined as child height for age below minus two standard deviations (−2SD) from the median height for age of the reference population.
- **Wasting:** Defined as child weight for height below minus two standard deviations (−2SD) from the median weight for height of the reference population.

## Methodology

### Study design and setting

CHAMP study is a population based, prospective cohort study involving mother-infant pairs (dyads) from District Swat, Pakistan. Swat is located at 34°46′58″ N and 72°21′43″ in Khyber Pakhtunkhwa province of Pakistan and home to more than 2.3 million people (31). Because of its geographic location in Hindukush–Himalayan region, majority of the land is occupied by mountains and thick forests. The area is one of the most marginalized and neglected regions of the province with livestock, agriculture, horticulture and tourism as the primary source of income and livelihood of the local population (32). Swat is also among the most vulnerable district in Pakistan that is prone to climate change. Since 2010, the district has been hit by at least three major floods that have significantly disrupted essential services such as healthcare, sanitation and access to safe drinking water (33). As a result, the majority of the population especially children are at high risk of malnutrition and its deleterious consequences.

Administratively, the district is divided into 07 administrative units know Tehsil Municipal Administration units (TMAs). Tehsil Matta has been chosen as the study area because it is the second most populated tehsil in district Swat with highest number of rural and remote communities. Following a preliminary survey, UC Beha was chosen as study site due to several reasons. It is located at high altitude with longest adjoining borders with district Dir (Upper) and most of the population is living in agropastoral communities with very limited access to education and healthcare facilities. In winters, access to the area is further limited by heavy snowfall and land sliding.

### Sampling technique

District swat is divided into 7 administrative units know as Tehsil Municipal Administration units (TMAs). Of these, tehsil Matta has been chosen as the study area because it is the second most populated tehsil in district Swat, located at high altitude with longest adjoining borders with district Dir (Upper) with highest number of rural and remote communities. The Matta tehsil is further divided into union ouncils, the primary administrative institutions in Pakistan. In rural areas, union councils are called village councils. Of the total 13 village councils in the sampling area, three union councils were randomly selected as the primary sampling unit. Subsequently, the researchers visited the primary health centers and designated vaccination centers within the chosen village councils. The newborn children were identified from the Extended Program for Immunization (EPI) registers. Potential participants were appraoched with the help of Lady Health Workers (LHWs), community based health workers linked with local health facilities. All the families in the area are registered with a designated LHW who visit them regularly. Important information such as the child’s name, father’s name, date of birth and address were recorded, and a list of eligible study participants was prepared. Once recruited, each participant will be followed for two years.

### Participants selection

The study population include 70 mother-infant pairs (dyads) according to the following criteria. Inclusion criteria

- Apparently healthy infants of both genders aged 0 – 28 days
- Born by natural or caesarean delivery
- Born to parents from district Swat
- Parents/caregivers have no plans to move out of the stud site for at least two years after enrollment in the study.

The exclusion criteria are:

- Child born to an underage (<18 years old) mother.
- Infants born with severe acute or chronic medical conditions that require hospitalization or prolong use of medication or both or the infant is diagnosed with enteropathies.
- Weight of the child is <1500 gm.

Ethical approval of the study was granted by the Research Ethics Board of Khyber Medical University. The study is also registered on www.clinicaltrial.gov (NCT05793294

### Recruitment

In local (Pashtun) culture, it is not possible to directly contact a potential female study participant without prior consent from the husband/male head of the household. Therefore, at each study site, an initial information session was held with the male parents/guardians with the help of local, elected representatives and elders of the community. During the session, complete information about the study objectives, eligibility criteria and data collection process were presented and explained by the team leader in the local (Pashto) language. A participant information sheet containing all the study details and procedures in an easy to understand, local (Pashto) and Urdu (national language of Pakistan) were also provided to the parents during the session. All queries were also answered. Following the session, eligible parents/guardians were invited by trained research assistants to enroll their baby into the study. Once agreed, the parents were asked to sign a written informed consent form in their preferred language.

### Patient and public involvement

The community members or the participant’s parents were not directly involved in developing study design, conduct, and outcome measures. The study protocol was reviewed and approved the Office of Research, Innovation and Commercialization (ORIC) Khyber Medical University and National Institute of Health (NIH) Pakistan. Healthcare professionals (LHWs) involved in the study have close liaison with the community and act as bridge between the researchers and participants. They also contributed in designing data collection questionnaires, informed consent and helped ensuring confidentiality. However, individual data will not be reported back to the participants.

### Study procedures

Figure 1 gives an overview of the overall study flow and study

**Figure 1:**
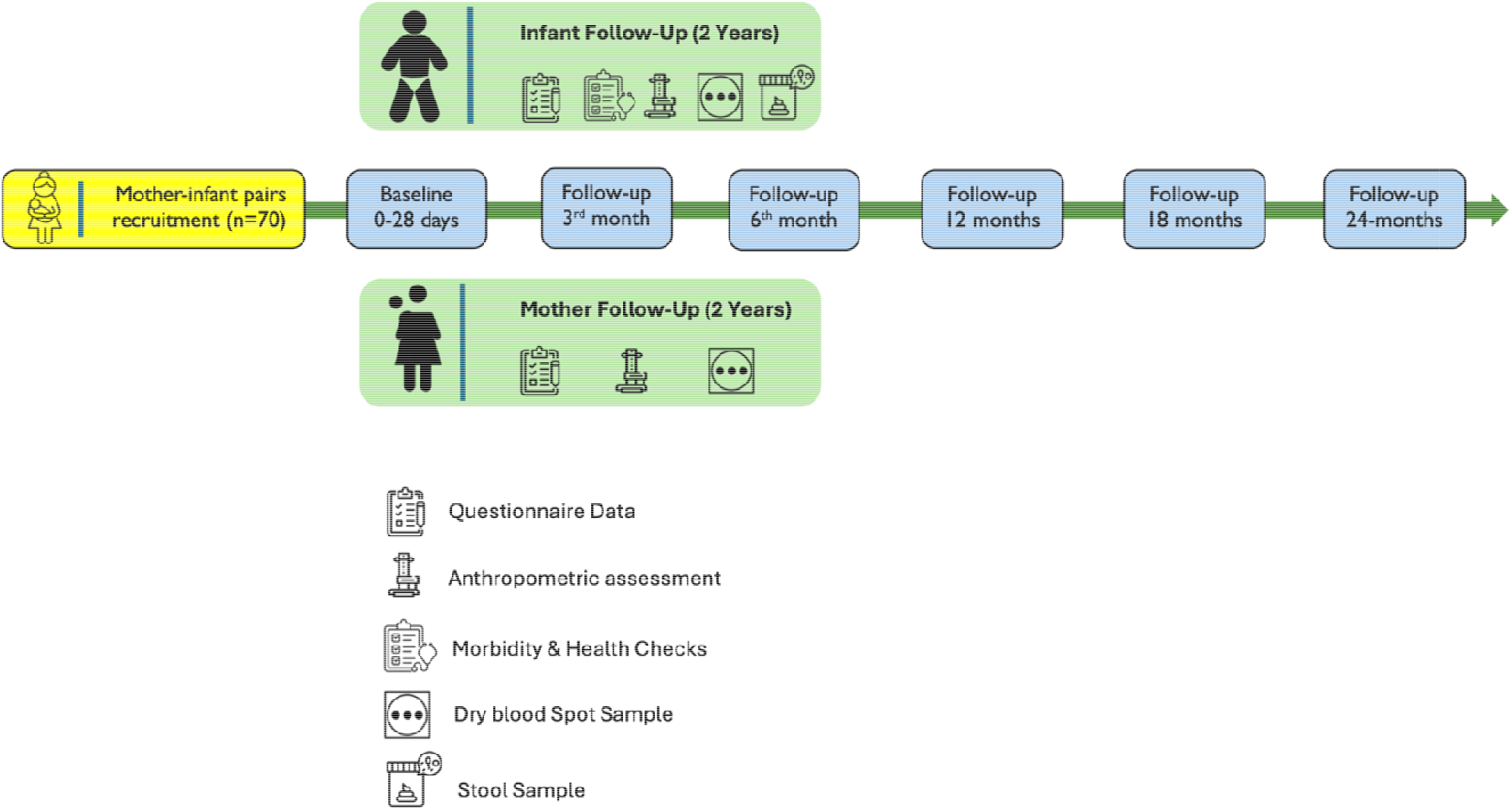
Flowchart of the study

### Data collection

We will collect different data from both the mother and infant at baseline and each time point during the follow-up period. The data collection in this project used an Android version of the KoBoCollect/Toolbox as a data collection instrument. To collect demographic and socioeconomic information of the study participants, an interviewer administered, structured questionnaire will be used. The questionnaire includes information such as name age, gender, household information and socio-economic status. Details about ante-natal and post-natal care will also be recorded using a structured, validated questionnaire adapted from the National Nutrition Survey Pakistan questionnaire (34). These data were recorded only at the baseline, i.e. start of the study.

Dietary Quality Questionnaire (DQQ) was used to assess dietary intake of the mothers. The method has been chosen because in Pakistan, there is no validated food frequency questionnaire to assess dietary intake. DQQ is a standardized and easy to use tool to collect food group consumption data required for calculating diet quality indicators such as Minimum Dietary Diversity for Women (MDD-W). The questionnaire has been validated and implemented across more than 100 countries including Pakistan (35). DQQ is a list-based method consisting of yes/no responses to 29 food groups consumed by the respondent during the previous day and night (24 hours). The procedure typically requires around 5 - 7 minutes to complete. Information regarding feeding practices of the children will be collected using validated, semi-structured questionnaire based on Infant and Young Child Feeding Practices (IYCF) indicators developed by Technical Expert Advisory Group on Nutrition Monitoring (TEAM) of WHO (36). This is a set of simple, validated and reliable indicators, widely used to assess IYCF practices across the globe especially in low- and middle-income countries. IYCF indicators are assessed by interviewing mothers or the main caregivers. The mothers will also be interviewed to collect data about child medical history, diarrhea and respiratory tract infection, hospitalization and use of antibiotics using a structured questionnaire. DQQ, IYCF and child health record data will be collected at baseline and at each time point during the follow-up.

### Anthropometric assessment

At baseline and each time point, the following anthropometric assessment will be performed on each child/infant and their mothers, using the standard methods (37).

#### Height/length

Recumbent length will be measured using an infantometer. Before measuring, the measuring board will be placed on a hard, flat surface such as a ground or sturdy table. The mother was asked to place the child gently on the board such that the infant’s head is aligned against the headboard while an assistant straightens the infant body and feet. When the child is position is correct, the researcher moves the foot board firmly against the child’s heels. The length of the child is recorded to the nearest 0.1 cm. The measurement will be repeated twice, and the average of the two measurements is recorded as the final length of the child. Mothers’ height will be recorded using a stadiometer following standard protocols. Briefly, they will be asked to remove shoes and gently stand on stadiometer. To ensure accurate measurement, the mothers will be instructed to stand up against the board, ensuring the Frankfurt plan position in which the back of the head, shoulder blades and buttocks and heels touch the back of the stadiometer. Once ready, the researcher will gently and firmly slide the measuring board’s moveable headpiece down until it touches the crown of the person’s head (compresses the hair). The procedure is repeated twice and the average of the two measurements is recorded as the height of the mother.

#### Weight

To record the weight of the child, a specialized electronic scale (Secca, UK) will be used. Before weighing the child, the parent will be instructed/helped to remove shoes, socks and heavy clothing except dry clean diapers or underpants from the child. When ready, the child is placed on his/her back on the center of the scale pan with the help of a trained researcher. We wait until the infant is still in the pan and the digital display is no more changing. The weight of the child is recorded to the nearest 0.01 kg. The weight of the mother is measured in kilograms using a calibrated electronic weighing scale. The mothers will be asked to remove shoes, jewelry and extra clothing (dupatta/chaddar) and stand calmly on the scale. The weight will be recorded to the nearest 0.01 kg twice and the average of the two measurement is considered the final weight of the mother.

#### Mid upper arm circumference (MUAC)

For infant MUAC measurements, a non-stretchable, numbered and colored MUAC tape will be used. To prepare for the measurement, the infant mother will be asked to sit on a comfortable chair, place the child in her lap and remove any clothing covering the left hand of the child. The mid-point between the tip of the elbow and the shoulder is located by straightening the arm along the axis of the body and forearm at 90-degree forwards. Once the mid-point is marked with a pen, the arm is straightened and the MUAC tape is placed around the arm at the midpoint and the reading is taken to the nearest mm.

#### Head circumference

The head circumference of the child will be measured using a non-stretchable measuring tap around the most prominent part of head to the middle of the forehead while the hair and soft tissue is compressed. At least two measurements will be recorded and the average of the two will be taken.

### Sample collection

#### Dry blood spot

Dry blood spots (DBS) are collected from both mother and the infant at baseline and during the follow-up by skin puncture of the third or fourth finger of the non-writing hand (38). Before DBS collection, the participant’s hands are first warmed followed by anterograde massaging of the finger in the direction of the blood flow towards the puncture site. The DBS collection site i.e. palmar side of the tip distal phalanx is cleaned with 70% isopropyl alcohol and punctured using a single use lacet. The first drop of the blood is wiped off with a gauze pad and subsequent drops are transferred to the marked circles on the surface of the filter paper without touching the surface. After drying, the DBS papers are transferred to a zip lock bag and stored at -80 °C till further processing in KMU main lab.

#### Stool

Stool samples are collected from the diapers of the infants at each time point. For this purpose, the parents are provided diapers, a specimen collection jar, disposable gloves, a zip lock bag and an instruction sheet, one day before the sample collection. The parents are instructed to regularly check their child’s diaper and remove it immediately after the child passes the stool. Stool samples are transferred immediately to the specimen container, put in a zip lock bag and handed over to the research assistant within 4 hours of the sample collection. The research assistant transfers 400 mg of stool sample into 2 mL screw top tubes prefilled with DNA shield reagent (Zymo Research USA) and sends it to main KMU lab at room temperature where they will be stored at -80 °C.

### Laboratory analysis

#### Hematological aassessment

Dry blood samples will be used to assess biomarkers of infections, micronutirents deficiencies and inflammation. For this purpose, commercially available, chemiluminscence based Q-plex aray kits (Quansys Biosciences) will be used following manufacturer instruction. This assay reliably detect and quantify histidine-rich protein II (biomarker of malarial infections), C-reactive protein, alpha-1-acid glycoprotein (biomarkers of inflammation), ferritin and soluble transferrin receptor (biomarker of iron deficiency), retinol binding protein (biomakrer of vitamin A deficiency) and thyroglobulin (biomarker of iodine deficiency) (Qu (39).

#### DNA extraction and 16S rRNA sequencing

Bacterial genomic DNA will be extracted from fecal samples (200 mg) stored at -80 °C using a ZymoBIOMICS DNA Miniprep Kit (Zymo Research, Irvine, CA, USA) following the manufacturer’s instructions. Each sample will be quantified using a Denovix and quality-checked through PCR of the 16S rRNA gene target and agarose gel electrophoresis. For the amplicon PCR step, 12.5 ng of purified bacterial genomic DNA will be used to amplify the V3-V4 target region according to the manufacturer’s protocol. The amplified samples will then be subjected to agarose gel electrophoresis to separate the DNA samples from primer dimers. Each DNA band will then be extracted and purified using a QIAquick gel extraction kit (Cat. no. 28705 QIAGEN® Germany). After purification, the concentration of each amplicon PCR will be quantified by Nanodrop. Next, the library preparation will be performed via indexing PCR and the reaction subsequently cleaned up and quantified, followong Illumina kit’s guidelines. Concentrations of all samples will then be standardized to a minimum of 2.5 nmol. One µL of DNA from each of these samples will then be pooled together and sent for high throughput sequencing on the Illumina Miseq® (Illumina, San Diego, California, USA) platform following the manufacturer’s instruction.

#### Monitoring and evaluration stretegy

Monitoring and evaluation of the whole project will be in place at all levels. Data entry will be randomly rechecked for accuracy by re-interview or re-visit to households taking part in the study. Principal investigator will oversee the data collectors and data enumerators. Bimonthly assessments will be done by the project team consisting of the project manager, PI and co-PI. In addition, KMU ORIC team will monitor the overall progress of the project through their own monitoring and evaluation system. Any discrepancy or deviance from the main objectives or lapse in the time frame and data collection procedures will be handled in close coordination of the project team with the filed data collector and support from KMU.

#### Participants retention strategies

A common issue with longitudinal cohort studies is systemic attrition which can have significant impact on generalizability of outcome of interest. Study participants may lose interest in the study and therefore decline participation especially in long duration cohort studies. Below is a list of potential reasons for attrition in the current project and mitigation strategies.

1. The first and most important reason might be that some participants may leave the study district permanently and leaving the research team unable to track and collect data at different time points. In order to tackle this issue, we have set our inclusion criteria such that only those participants who are permanent residents of this districts and who confirm that they have no plans to move outside of the district permanently, will be recruited.
2. Reducing barriers to participation by hiring local research assistants and their training, ensuring anonymity, translating the data collection tools to local language (Pashto) and pilot testing.
3. Setting up a follow up and reminder strategy by calling the participants before the data collection time.
4. Tracing via alternative phone numbers and visiting the house physically when the participants are not responding.

## Data analysis

All *in sillico* data processing and analysis will be performed on IBU Linux Cluster for Rocky Linux 8 from the servers of the University of Bern, on a local machine running Ubuntu 20.04.06 LTS (32GB, Intel® Core™ i7-9750H CPU @ 2.60GHz × 12) and within R (4.4.1 2024-06-14) package environments in RStudio (2024.04.2). Sequence reads obtained for each sample will be processed for quality with FastQC (v0.11.9) and MultiQC (version 1.11); further quality checks, reads trimming, chimeras removal, denoising and taxonomy assignment (using a trained Naive Bayes kmer classifier) will be performed directly from within QIIME2 workflow (q2cli version 2024.5.0) using the included DADA2 software.. Standard core metrics such as alpha diversity indices (Faith’s richness, Simpson’s evenness, and Shannon index) and beta diversity (Bray-Curtis, weighted unifrac for the quantitative ones; jaccard and unweighted unifrac for the qualitative ones; Aitchison) will be computed with QIIME2, then explored in Qiime2View and further processed and compared with Phyloseq and qiime2R packages in R. We will use the Emperor biplot plugin, Principal Coordinate Analysis (PCoA) and beta-group-significance plugin to test beta diversity significances and to characterize the top bacterial genera that explain the microbiota distributions across the samples.

We will seek to identify any correlations between the collected metadata and microbiome composition, and identify any gut/oral microbiota trends associated with nutritional status and food diversity. We will decorticate the variations in the observed microbiota composition across different groups, e.g Weight, Height, Age, Nutrional status. For alpha diversity comparisons, the groups’ significances will be computed and tested with Kruskal-Wallis pairwise tests. We will also use the q2-longitudinal plugin to investigate longitudinal and paired sample comparisons, i.e to determine if/how samples change between the different time points and treatments/nutritions. All the post-processing and statistical analysis will be performed in R. Weight for height, weight for age, height for age z-score will be calculated using WHO Anthroplus software and compared with microbiome composition using R.

### Status of the study

The study start in my 2024 and will continue till May 2026. As of October 2024, 70/70 (100%) participants have been recruited. The data and samples have been collected for the baseline (n=70) and first follow up period (3 – months; n = 66).

## Discussion

The CHAMP study aims to longitudinally characterise gut microbiome development in newborn infants during the first two years of life and includes 70 mother-infant pairs (dyads) recruited from remote, rural communities of District Swat, Pakistan. The study involves comprehensive data collection regarding dietary intake, nutritional status, infant growth and morbidities at different time points. Besides, prospective collection of stool samples will allow gut microbiome assessment over time and identify demographic, mother and child related factors affecting its diversity and abundance. The study findings will contribute towards better understanding of the gut microbiome colonization and development, associated factors and its impact of on growth in infants from malnutrition endemic area of Pakistan.

Microbial colonization of the gut during infancy plays a crucial role in establishment of intestinal barrier and development of immune system (40). Numerous studies have reported rapid changes in the composition and diversity of microbiota during first two years of life in healthy infants. However, the pattern and extent of microbial colonization is largely influence by the feeding practices and nutritional status of the infants. There exists a bidirectional relationship between gut microbiome development and nutritional status. Malnutrition can impact gut microbiome development and maturation during early childhood resulting in dysbiosis. As a result, an immature and altered gut microbiome leads to malnutrition due to impaired energy production, vitamin biosynthesis and immune dysfunctions (41,42). These findings are critical especially in context of developing countries like Pakistan where childhood malnutrition is a public health issue (34). The CHAMP study will help to dissect the relationships between dietary intake, nutritional status and gut microbiome phenotypes during infancy and pave way for developing microbiome-based interventions to tackle the problem of malnutrition in Pakistan.

Our study is the first of its kind to characterize gut microbiome development in newborn infants from malnutrition endemic area of Pakistan. The study will greatly help to understand the infant growth trajectory in relation to gut microbiome development and provide conceptual basis for sustainable nutrition recommendation and interventions in future. The prospective longitudinal nature of the study will help collection of detailed data with minimal constraints (6 visits in 2 years) and associated risks (no intervention and minimally invasive procedures to collect biological samples). The study will obtain precise data on dietary intake and nutritional status and their short term and medium-term impacts on gut microbiome development and vice versa. To get deeper insights into how these factors especially dietary intake pattrens and socioconomic status impact gut microbiome development, we will compare our study results with similar studies conducted in Zimbabwe, a low income country (The University of Zimbabwe College of Health Sciences (UZ-CHS) Birht Cohort) (43) and Switzerland, a high income country (The Bern Birth Cohort – BeBiCo study) (44). Both these studies involve mother infant pairs and collect nealry idential data on demographics, socioeconomic status and diet at time points corresponding to our study. These studies also collect fecal samples which will allow us to compare gut microbiome development in Pakistani infants with infants from Switzerland and Zimbabwe, the countries with completely different landscapes in terms of environmental, culture and dietary intake patterns.

Our study also has some limitations. First, the launching and implementation of a cohort study in remote, difficult to access, rural areas are challenging, anywhere in the world including Pakistan. As a result, the drop out and attrition in sample size is expected. Second, study involves multiple assessments requiring the mother to travel long distance in mountainous areas to reach the sampling site. The process is time consuming and potentially burdening on mother and infants. However, these issues will be addressed partly by providing compensation for travel and sending advance reminder about follow-up visits using phone calls. Third, due to self-reported and retrospective nature of the data collection during each visit, the response and social desirability bias cannot be avoided fully. Finally, we can only identify the association between growth and gut microbiome development but no causal relationship because of the non-interventional study design.

## Data Availability

All data produced in the present study are available upon reasonable request to the authors

